# RIGHT SEPSIS CLASSIFICATION- MUST FOR ANTIMICROBIAL STEWARDSHIP: A LONGITUDINAL OBSERVATIONAL STUDY

**DOI:** 10.1101/2024.08.07.24311603

**Authors:** Jaideep Pilania, Prasan Kumar Panda, Ananya Das, Udit Chauhan, Ravi Kant

**Affiliations:** Department of Internal Medicine, All India Institute of Medical Sciences (AIIMS), Rishikesh; Department of Radiology, All India Institute of Medical Sciences (AIIMS), Rishikesh

## Abstract

**Background:** Sepsis is a critical medical condition characterized by life-threatening organ dysfunction triggered by a dysregulated response to infection. It poses a substantial global health burden, with significant morbidity, mortality, and economic costs, particularly pronounced in low- and middle-income countries. Effective management of sepsis relies on early recognition and appropriate intervention, underscoring the importance of accurate classification to guide treatment decisions.

**Objective:** This longitudinal observational study aimed to assess the distribution of sepsis categories and the use of empirical antibiotics classified by the WHO AWaRe system in a tertiary care hospital in Northern India. The study also aimed to highlight implications for antimicrobial stewardship by examining the use of AWaRe group antibiotics and their correlation with sepsis classifications.

**Methods:** A total of 1867 patients admitted with suspected sepsis were screened, with 230 meeting inclusion criteria. Patients were categorized into different sepsis classes (Asepsis, Possible Sepsis, Probable Sepsis, Confirm Sepsis) and followed until discharge or Day-28. Descriptive statistical analysis was employed to assess sepsis categories and empirical antibiotic usage classified by Access, Watch, and Reserve categories according to the WHO AWaRe system.

**Results:** Among the study cohort (mean age 40.70 ± 14.49 years, 50.9% female), initial sepsis classification predominantly included Probable Sepsis (51.3%) and Possible Sepsis (35.7%), evolving to Asepsis (57.8%) upon final classification. Empirical antibiotic use showed a concerning predominance of Watch group antibiotics (92.5%), with Ceftriaxone (45.7%) and piperacillin-tazobactam (31.7%) being the most commonly prescribed.

**Conclusion:** The dynamic nature of sepsis classification underscores the complexity of diagnosing and managing this condition. Accurate categorization is pivotal for clinical decision-making, optimizing antibiotic use, and combating antimicrobial resistance. The majority of the asepsis category was levelled as probable or possible sepsis and given antibiotics. The high reliance on Watch group antibiotics in empirical therapy signals a need for enhanced diagnostic strategies to refine treatment initiation, potentially reducing unnecessary antibiotic exposure. Future efforts should focus on establishing sepsis classification checklists and promoting adherence to antimicrobial stewardship principles to mitigate the global threat of antimicrobial resistance.

## INTRODUCTION

### Background/Rationale

Sepsis, a life-threatening condition, characterized by life-threatening organ dysfunction triggered by a dysregulated response to infection, has become a growing concern in the medical community due to its significant impact on patient outcomes and healthcare costs. Early identification and prompt intervention are critical in the management of sepsis, as the condition is time-sensitive and can rapidly progress to organ dysfunction and shock.

Sepsis is a syndrome which has physiologic, biochemical and pathologic abnormalities which are induced by the infection, which accounts to more than $20 billion of the total United States hospital costs, as per 2011 data (1). As per The Global Burden of Disease study, it is was estimated that there were around 48.9 million sepsis cases worldwide in the year 2017, with around 11 million deaths related to sepsis in the same year, which accounts for around 19.7% of all global death (2). The majority of sepsis related burden is found in the low- and middle-income countries (LMIC) in the world. In various developing countries, such as India, with a population of more than 1.4 billion people, the data regarding prevalence, apart from other epidemiological data, is missing and poorly understood, despite high rates of mortality related to sepsis. In the year 2017, the estimated cases of sepsis in India were around 11.3 million, and among them 2.9 million deaths were noted (3). To know the latest epidemiological data regarding sepsis, a study named “Sepsis in India Prevalence Study (SIPS)” is ongoing.

As per the “Third International Consensus Definitions for Sepsis and Septic Shock (Sepsis-3)” guidelines, sepsis has been defined as life-threatening organ dysfunction which is caused by a dysregulated host response to infection (4). Over the time, the definition of sepsis has undergone various changes. There are various scoring systems available for assessing the mortality and outcomes in patients with sepsis. Sepsis is an extremely heterogenous entity, both at clinical and molecular levels, which can have various subtypes and there are even conditions which can mimic sepsis, i.e sepsis mimics (5). For better, prompt and standardised management of sepsis, the concept of bundle care was made, which has also undergone many changes with time.

There is growing incidence of antimicrobial resistance all around the global and it is emerging as a public health concern, and has led to emergence of many multidrug resistance microbes, also called as “superbugs”. It needs to be taken care with utmost priority due to decreasing number of available options of antibiotics for treatment of patients with such multidrug resistance organisms, leading to increase in morbidity and mortality. In the year of 2017, WHO Expert Committee on Selection and Use of Essential Medicine develop the AWaRe classification of antibiotics as a tool which can support antibiotics stewardship at various levels (6). The antibiotics were categorized into 3 groups-Access, Watch and Reserve on the basis of their impact on the antimicrobial resistance (AMR). This action is a step forward to reduce the burden of antimicrobial resistance (AMR) by following the targets set by WHO. To understand the importance of this, it is to be noted that in the year 2019, it was estimated that AMR was responsible for around 1.3 million people’s death worldwide (7).

Inappropriate use of antibiotics in humans is the major well-known and proven factor for antimicrobial resistance (AMR), apart from other causes (8). The misuse of antibiotics, including using them unnecessarily or selecting the incorrect antibiotic at the wrong dose, for the wrong length of time, and through the wrong method, is a prevalent issue affecting 30% to 50% of all antibiotic prescriptions (9,10).

### Objective of Study

This longitudinal observational study was done at a tertiary care teaching hospital in northern India with the objective to assess the distribution of various sepsis categories (asepsis, possible sepsis, probable sepsis, and confirm sepsis) and the use of empirical antibiotics as per WHO AWaRe classification. The study also aimed to highlight implications for antimicrobial stewardship by examining the use of AWaRe group antibiotics and their correlation with sepsis classifications.

## METHODS

### Study Design

This was a longitudinal observational study

### Study Setting

Study was done at a tertiary care teaching hospital in northern India from 1^st^ January to 31^st^ December, 2023 in the department of general medicine, after the approval from Institute Ethics Committee (IEC). Data from patients was collected and entered in RedCap software (AIIMS Rishikesh version), and also in the Microsoft Excel sheet.

### Objective

-To assess the distribution of various sepsis categories (asepsis, possible sepsis, probable sepsis, and confirm sepsis).

-To assess the use of empirical antibiotics as per WHO AWaRe classification.

### Participants

Participants include patients who were admitted in the department of general medicine, and were eligible as per inclusion and exclusion criteria of the study between 1^st^ January to 31^st^ December, 2023.

### Inclusion Criteria

**-**Patients of age >/= 18 years who are admitted in department of general medicine with suspected sepsis.

### Exclusion Criteria

-Patients who were diagnosed etiologically other than sepsis with 5 days of hospital admission

-Patients whose data was missing.

### Variables/ Outcomes

-To estimate proportion of patients in different categories of sepsis at admission and outcome (death/discharge).

-To estimate empirical antibiotics use as per WHO AWaRe classification.

### Data source

Data was collected from patients admitted in general medicine ward, along with medical records of discharged patients.

### Study size

The study size or sample size was not mathematically calculated, as no prior refence study was available for same. So, as per feasibility, universal sampling method was used for samples of the study.

### Statistical Method

The study was primarily an observational study which used descriptive data analysis. Patients with missing data were not included for the final result analysis.

## METHODOLOGY

-Patients admitted in Department of General Medicine with suspected sepsis were screened for inclusion and exclusion criteria of the study and patients fulfilling criteria were included in the study.

-Patients, both-admitted and discharged, were assessed for inclusion in the study. For discharged patients, data was extracted from available hospital records and then were subjected to 3 step approach/ model.

-A 3-step model was prepared using the sepsis definition, and were divided into various steps. Initial step included evidence of dysregulated host response, which was evaluated with the use of National Early Warning Score-2 (NEWS-2 score). As per the published report by Royal College of Physicians (RCP), a NEWS-2 score of 5 or more should make one think for sepsis [15]; thus, for our study, we took a higher value of NEWS-2 score as an evidence of dysregulated host response i.e a NEWS-2 score of ≥6 was used.

- “Suspected sepsis” term was used, which was defined by one the following parameters:

i. Need of antibiotics for management
ii. Evidence of infection anywhere in the body
iii. Organ dysfunction not explained by non-infective cause
iv. Patient improved after antibiotics.

The term of suspected sepsis, with any one of the above-mentioned points, was first validated by experts from various fields including internal medicine, infectious disease, etc, including both, from the institute experts and also not related to institute, and was then incorporated into the study.

-After screening and inclusion, patients were subjected to 3 step approach/ model and were followed, either physical or via available hospital data or records, till an outcome (which is discharge/ death) is reached, and were categorized into different sepsis categories on different day on follow-up.

-All baseline data was collected including vitals (for calculation of NEWS-2 score), laboratory data, cultures, and also the empirically used antibiotics.

- 3-step approach/ model has step-1 as evidence of dysregulated host response (assessed by the use of NEWS-2 score), step-2 was risk factor of infection and final, step-3, was to look for evidence of infection.

To identify and classify sepsis in a patient using novel 3-step model:

### STEP-1 EVIDENCE OF DYSREGULATED HOST RESPONSE

- NEWS-2 >/=6 was used as an evidence of dysregulated host response.

### STEP-2 RISK FACTOR FOR INFECTION

- Following were some of the most important risk factors for infection which were considered:

i. **Chronic illness/ co-morbidities** like-Decompensated Chronic Liver Disease (DCLD), Dialysis-dependent CKD, Uncontrolled DM (HbA1c ≥10%), Chronic lung diseases (Severe asthma/ Chronic Obstructive Pulmonary Disease/ Pulmonary Tuberculosis) etc.
ii. **Malnutrition** (BMI</= 18.5 kg/m^2^) like protein energy malnutrition (marasmus/ kwashiorkor)
iii. **Unhygienic living conditions** involving contaminated food & water supply (Risk factor for Hepatitis-A/ E, Typhoid, Cholera, Dysentery-bacillary/ amoebic, etc.)
iv. **Immunosuppressive states** like cancer, leukaemia, Nephrotic Syndrome, Long-term Steroid therapy, Febrile Neutropenia etc
v. **Age**-like Reproductive Females (Urinary Tract Infection/ UTI), Elderly (age>65 years).
vi. **Trauma/ wounds**
vii. **Structural diseases** like congenital heart disease (chd), cystic fibrosis, bronchiectasis etc
viii. **History of any surgery in past 1 year**
ix. **Any visible foci of sepsis** like folliculitis, cellulitis, pustule etc
x. **Recent travel history/ exposure**
xi. **Animal bite/ exposure** (Urine, Faeces, Saliva, Blood with Human Blood or its ingestion)
xii. **Previous hospitalization in past 90 days & Covid-19 infection.**

### STEP-3 EVIDENCE OF INFECTION: -

#### 3(A) CLINICAL EVIDENCE-SYNDROMIC DIAGNOSIS [16]

i. **Pyelonephritis**- Characterised by flank pain, tenderness or both and fever associated with dysuria, urgency and frequency
ii. **Infective Endocarditis**- Based on Modified Duke’s Criteria
iii. **Intra-Abdominal Infections – Peritonitis** (Fever, Abdominal pain with guarding & rigidity)
iv. **Skin & Soft Tissue Infections**- Furunculosis, Carbuncle, Necrotising fasciitis.
v. **Suppurative Infections**
vi. **Meningitis** – Fever, Neck stiffness, Headache/ Vomiting
vii. **Meningoencephalitis**- Fever, Neck stiffness & alteration of sensorium/ Seizures
viii. **Cerebrospinal Fluid (CSF) Shunt Infections**- Fever with headache/ nausea/ lethargy, tenderness or erythema over the subcutaneous tunnel and symptoms of peritonitis/pleuritis in patients with ventriculoperitoneal/ ventriculopleural shunts.
ix. **Catheter-related bloodstream infection (CRBSI)-** Presence of Bloodstream Infection and demonstrating that the infection is related to the catheter.
x. **Central line associated bloodstream infection (CLABSI)**- Laboratory confirmed blood stream infection (BSI) where an eligible blood stream infection (BSI) organism is identified, and an eligible central line is present on the day/ day before the event
xi. **Catheter related-UTI (CA-UTI)**- (a) Patient has at least one of the following signs or symptoms compatible with UTI with no other identified source of infection (i). New onset fever (ii). Supra-pubic tenderness (iii). Costo-vertebral angle pain or tenderness, acute haematuria (iv). Urinary urgency, urinary frequency, dysuria and (b) Patient has a urine culture with 1000 colony forming units (cfu)/mL of ≥1 bacterial species in a single catheter urine specimen or in a midstream voided urine specimen from a patient whose urethral, suprapubic, or condom catheter has been removed within the previous 48 hrs.
xii. **Osteomyelitis**
xiii. **Abscess** (Liver/ Lung/ Brain/ Muscle)
xiv. **Pneumonia**- Fever > 38.3° C (101.0° F) for >2 days and lasting up to 14 days and having received no specific treatment for this current illness with antimalarials or antibiotics.

#### 3(B) SUPPORTIVE/ SUGGESTIVE EVIDENCE

**-(i) Imaging:** Showing evidence of infection like

- X-Ray (usually a chest X-ray)
- Ultrasonography (USG)
- Endoscopy
- Computed Tomography (CT-Scan)
- Magnetic Resonance Imaging (MRI)
- Positron Emission Tomography (PET) Scan

**-(ii) Biomarkers:** Detected from samples such as-

a. Blood- Leucocytosis, Procalcitonin, Beta-1,3 glucan, Galactomannan etc.
b. Urine
c. Other Fluids like cerebrospinal fluid (CSF), Ascitic fluid, Pleural fluid.

#### 3(C) CONFIRMATORY EVIDENCE

**-(i) Direct Visualisation:** via Eye/Open Method i.e without use of any instrument like Myiasis, Ectoparasites etc.

**(ii) Endoscopic Evidence/ Visualisation**

**(iii) Microscopy & Culture Growth and Sensitivity-** Blood, Urine, Endotracheal Tube (ET) Aspirate/ Bronchoalveolar Lavage (BAL), Catheter Tip, Wound, Swab culture, Biopsy Material, Sputum, Other fluids like cerebrospinal fluid (CSF), Pleural, Pericardial, Ascitic, Synovial etc.

**(iv) Polymerase Chain Reaction (PCR)/ Gene Detection Methods**

**(v) Immunological Methods** like immunochromatography (ICT), Chemiluminescence immunoassay (CLIA), Enzyme-linked immunosorbent assay (ELISA) and others.

## 3-STEP MODEL FOR SEPSIS

**Figure-1:**
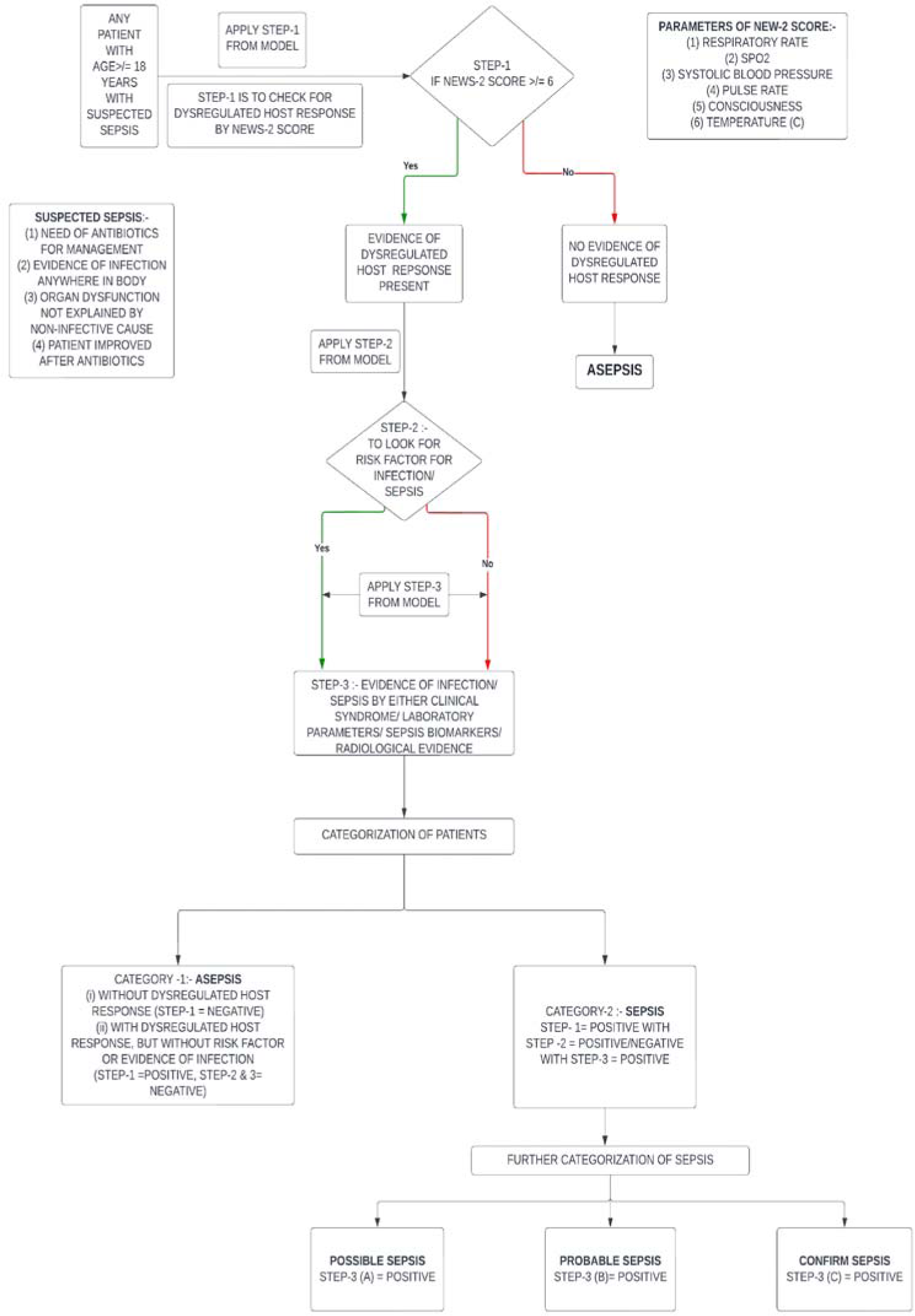
Flowchart to use the 3-Step model for sepsis classification.

## CATEGORIZATION OF SEPSIS

**Table-1:** Table showing various sepsis categories.

| **CATEGORY** | **INTERPRETATION** | **OUTCOME** |
| --- | --- | --- |
| CATEGORY-1 | (i) STEP-1 = NEGATIVE<br>(ii) STEP-1= POSITIVE WITH<br>STEP-2 & 3= NEGATIVE | **ASEPSIS** |
| CATEGORY-2 | STEP-1, 2 & 3(A) = POSITIVE | **POSSIBLE SEPSIS** |
| CATEGORY-3 | STEP-1, 2 & 3(B) = POSITIVE | **PROBABLE SEPSIS** |
| CATEGORY-4 | STEP-1, 2 & 3(C) = POSITIVE | **CONFIRM SEPSIS** |

## RESULTS

This longitudinal observational study was done which included patients aged >/= 18 years with suspected sepsis (inclusion criteria) who were admitted to the Department of General Medicine. A total of 1867 patients were screened and those diagnosed etiologically other than sepsis within 3 days of hospital admission were excluded from the study population, also excluding patients with missing data; resulting in a total study cohort of 230 patients, which were included for analysis which were followed till an outcome is reached.

The mean age (in years) was 40.70 ± 14.49 years for the analyzed participants. Of 230 participants, 113 (49.1%) were male and 117 (50.9%) were female, demonstrating slight female predominance [Table-2]. Age was further subcategorized into 3 major groups, and the majority (49.13%) belonged to 18-40 years of age, followed by 41-60 years (40.87%) [Table-2]. Demographic analysis revealed that the majority of participants were from 2 states, Uttar Pradesh (52.2%) and Uttarakhand (42.6%), attributed to the location of study tertiary centre, with other regions having their share of participants as shown in Table-2.

**Table-2:** Summary of Basic Details.

| Basic Details | Mean ± SD \|\| Median (IQR) \|\| Min-Max | OR | N (%) |
| --- | --- | --- | --- |
| Age (Years) | 40.70 ± 14.49 \|\| 41.00 (28.00-53.00) \|\| 18.00 - 70.00 |  |  |
| Age Group |  |  |  |
| 18-40 years |  |  | 113 (49.13%) |
| 41-60 years |  |  | 94 (40.87%) |
| >60 years |  |  | 23 (10%) |
| Gender |  |  |  |
| Male |  |  | 113 (49.1%) |
| Female |  |  | 117 (50.9%) |
| State |  |  |  |
| Uttar Pradesh |  |  | 120 (52.2%) |
| Uttarakhand |  |  | 98 (42.6%) |
| Bihar |  |  | 3 (1.3%) |
| Rajasthan |  |  | 2 (0.9%) |
| Delhi |  |  | 1 (0.4%) |
| Gujarat |  |  | 1 (0.4%) |
| Jharkhand |  |  | 1 (0.4%) |
| Madhya Pradesh |  |  | 1 (0.4%) |
| Punjab |  |  | 1 (0.4%) |
| Tamil Nadu |  |  | 1 (0.4%) |
| Telangana |  |  | 1 (0.4%) |

-Patients were classified into different categories of sepsis on the basis of devised novel sepsis classification system. On the basis of the classification system, the initial categorization was dominated by the majority from the probable sepsis (51.3%) and possible sepsis (35.7%) categories. However, during the final categorisation, the majority was formed by the asepsis group (57.8%), then followed by other categories. [Table-3].

**Table-3:**
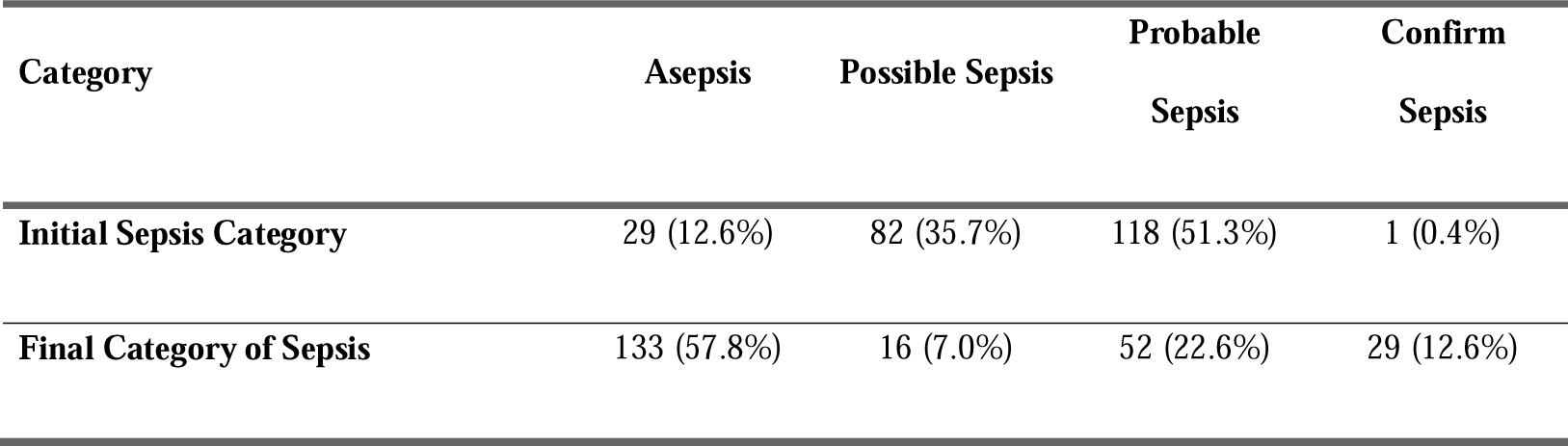
Summary of Sepsis Category.

-Figure-2 & 3 depicts the proportion of participants in different categories of sepsis on initial and final categorization using the novel classification system respectively. These figures also highlight the dominant category of sepsis at initial evaluation, probable sepsis, which, at the time of final evaluation was dominated by asepsis category.

**Figure-2.**
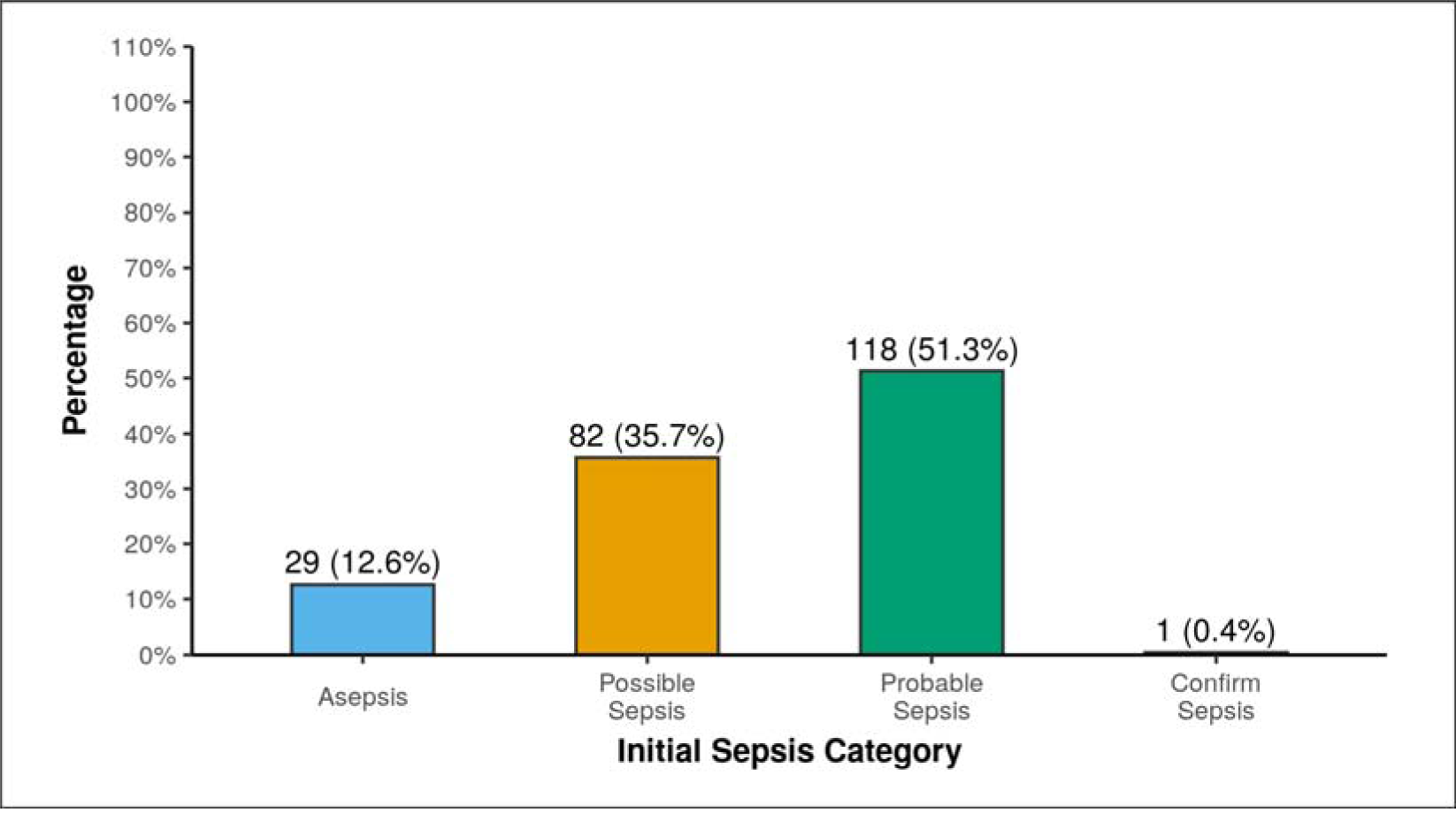
shows the proportion of patients in initial different categories of sepsis.

**Figure-3.**
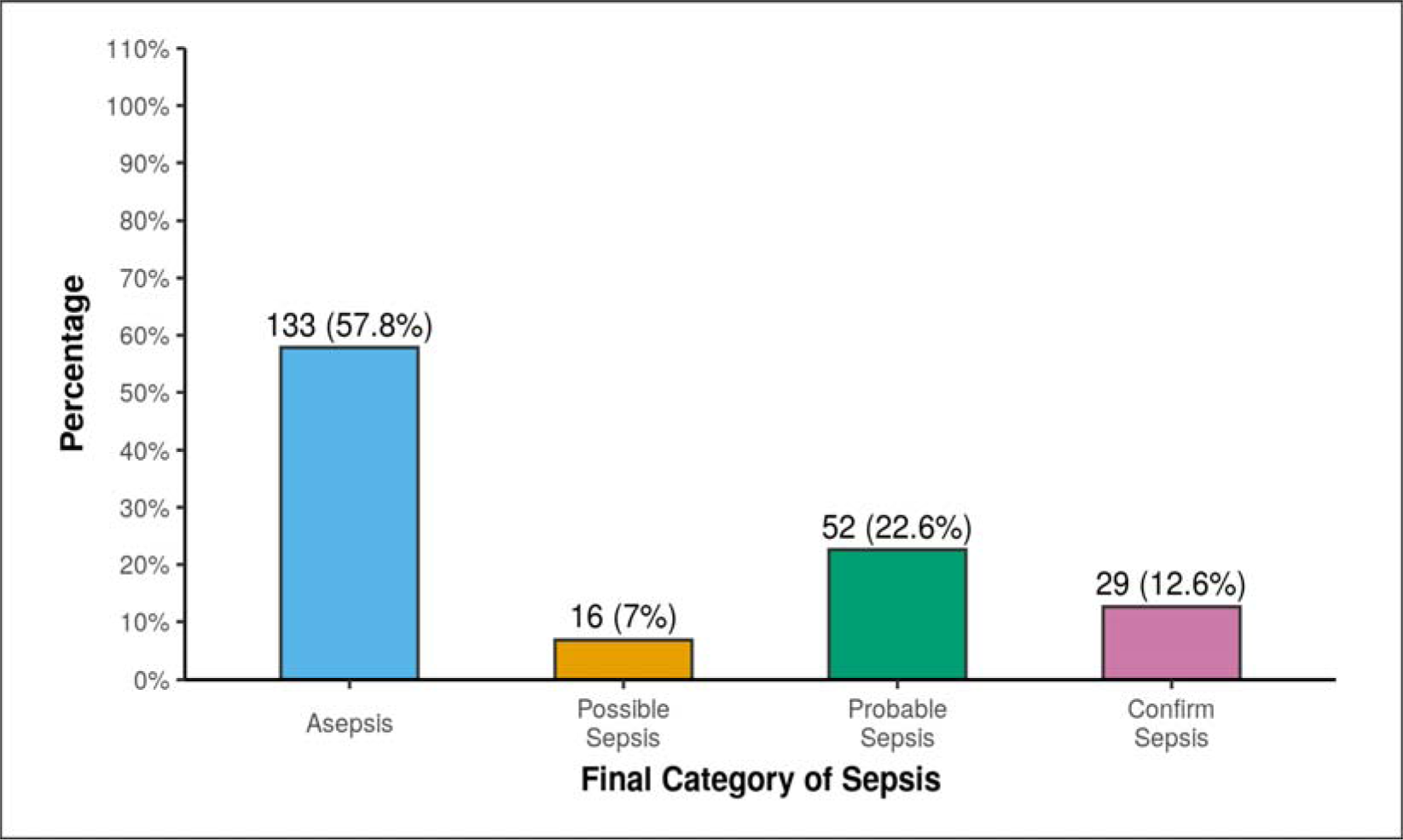
shows the proportion of patients in final different categories of sepsis.

-Figure-4 & 5 shows the percentage of patients in different categories of sepsis on both initial and final categorization. One can very easily notice the change in the sepsis categories during hospitalisation, with majority belonging to different category on different time intervals.

**Figure-4:**
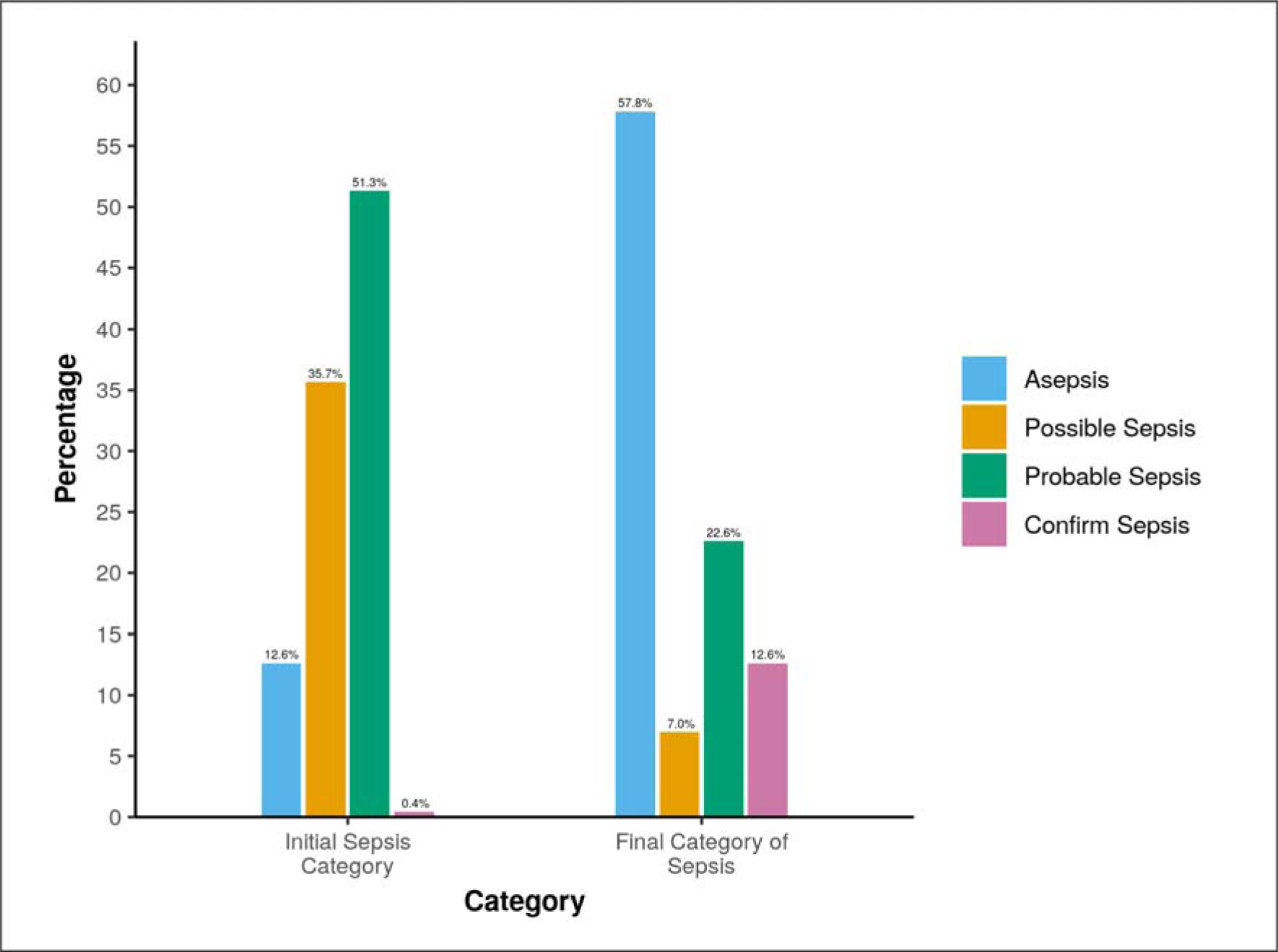
Graph showing percentage of patients in initial and final sepsis category.

**Figure-5:**
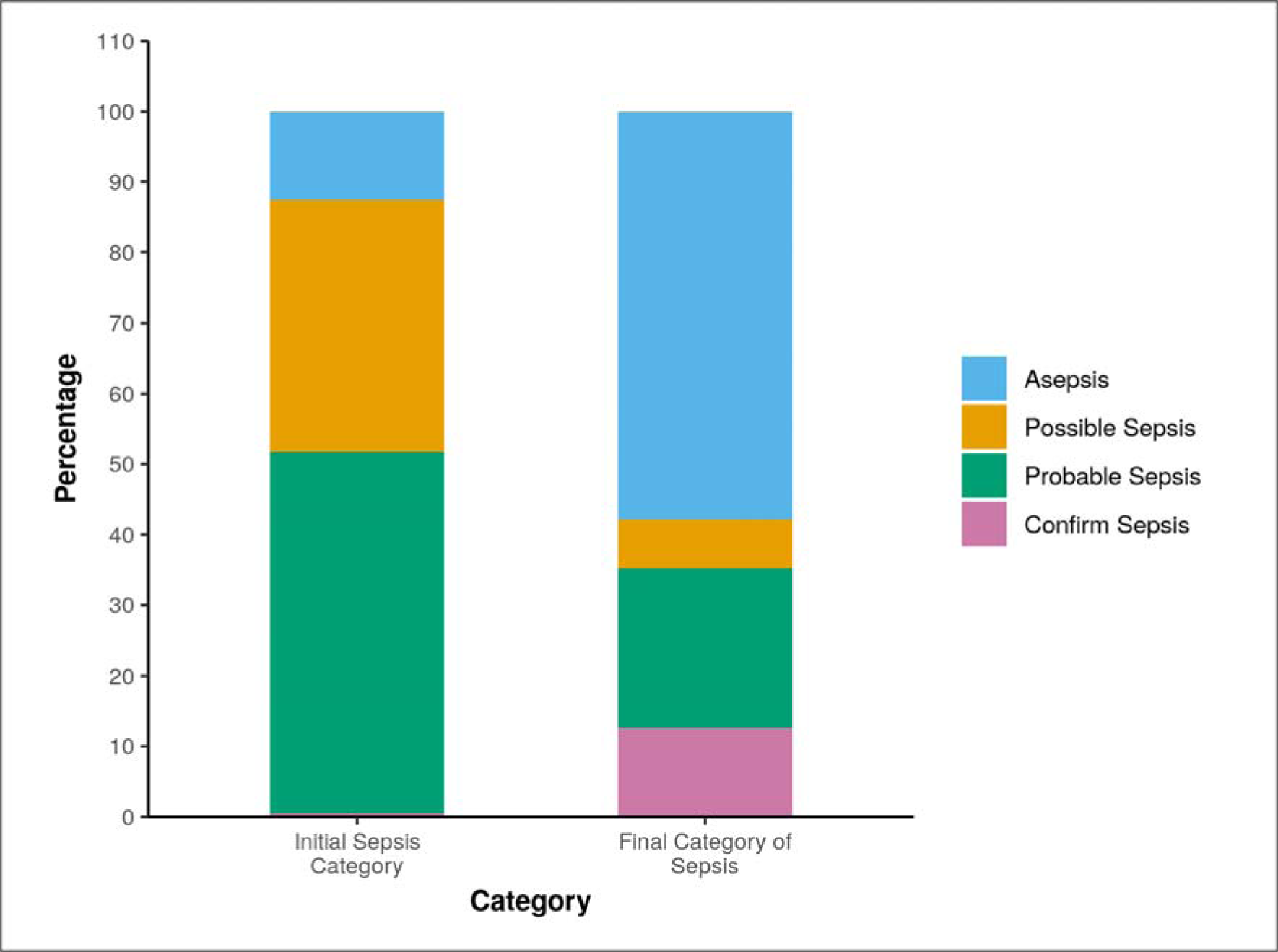
Graph showing proportion of patients in different categories of sepsis at various time points.

-To estimate the use of empirical antibiotics and to classify them as per WHO AWaRe classification system was also studied as an objective in this study. The results from the study revealed that the majority of empirical antibiotics used were from the ‘Watch’ group of AWaRe classification, accounting to more than 90% of patients receiving the ‘Watch’ group of empirical antibiotics, followed by ‘Access’ group [Table-4]. Similar findings are also depicted in graphical format in figure-6.

**Table-4:**
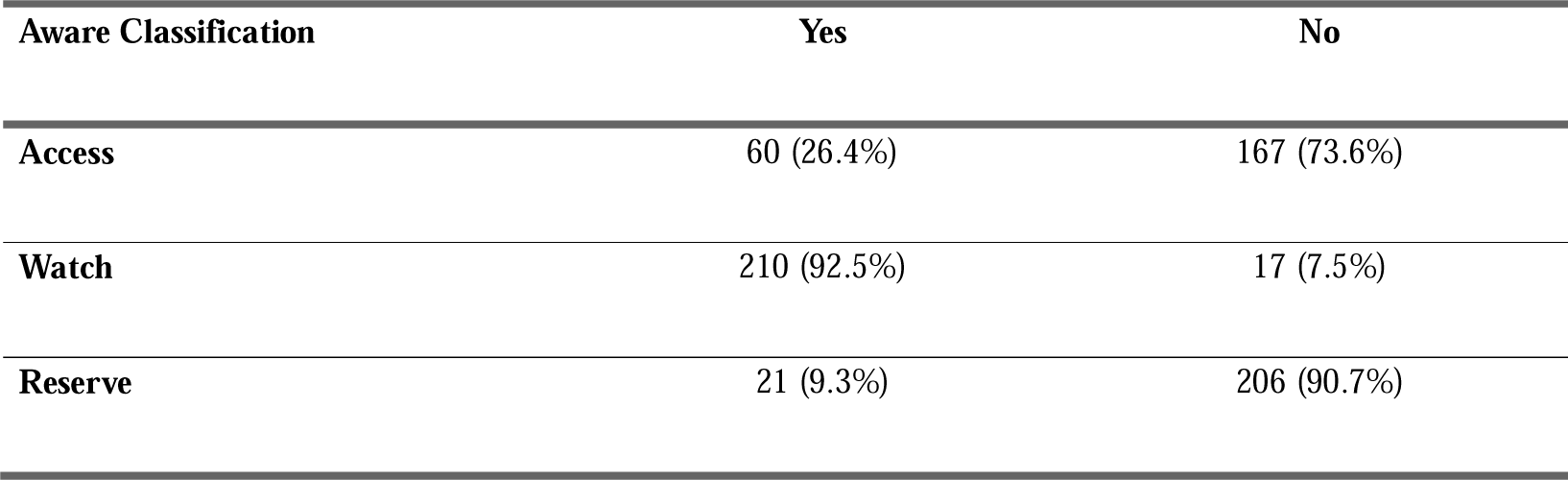
Summary of Aware Classification.

| Aware Classification | Yes | No |
| --- | --- | --- |
| Access | 60 (26.4%) | 167 (73.6%) |
| Watch | 210 (92.5%) | 17 (7.5%) |
| Reserve | 21 (9.3%) | 206 (90.7%) |

**Figure-6:**
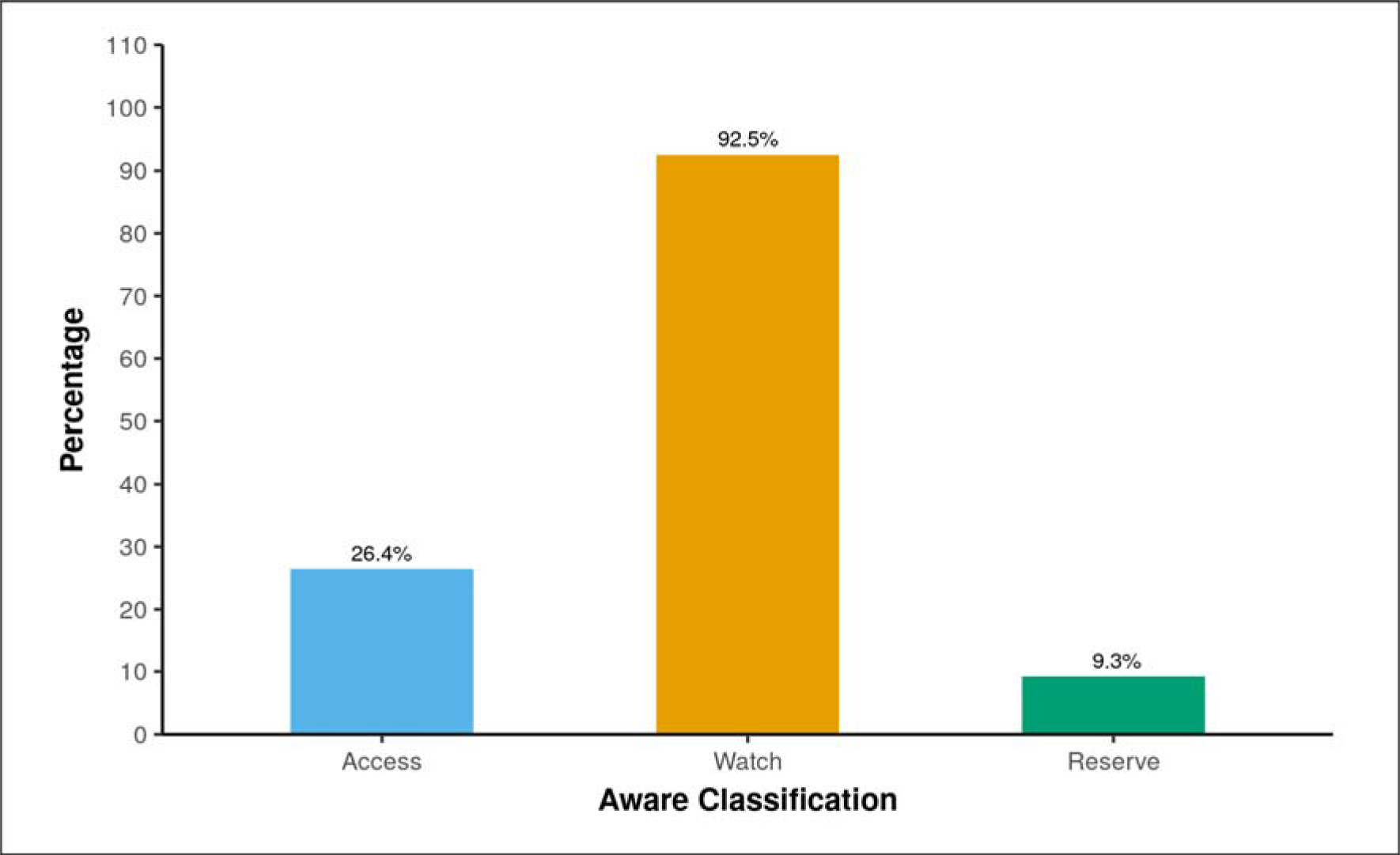
Graph showing the percentage of patients who received antibiotics as per AWaRe classification.

-Apart from the classification of empirical antibiotics as per WHO AWaRe classification, the study also found the proportion of various empirical antibiotics used, and found that the majority of patients received Ceftriaxone (injectable 3^rd^ generation cephalosporin) (45.7%), which was followed by piperacillin-tazobactum (injectable beta-lactum with beta-lactamase inhibitor) (31.7%), vancomycin (injectable glycopeptide) (22.6%) and azithromycin (oral & injectable macrolide) (22.2%) as empirical antibiotics [Table-5]. Figure-7 shows the graph that represents the percentage of empirically used antibiotics, representing the similar findings as presented in Table-5.

**Table-5:**
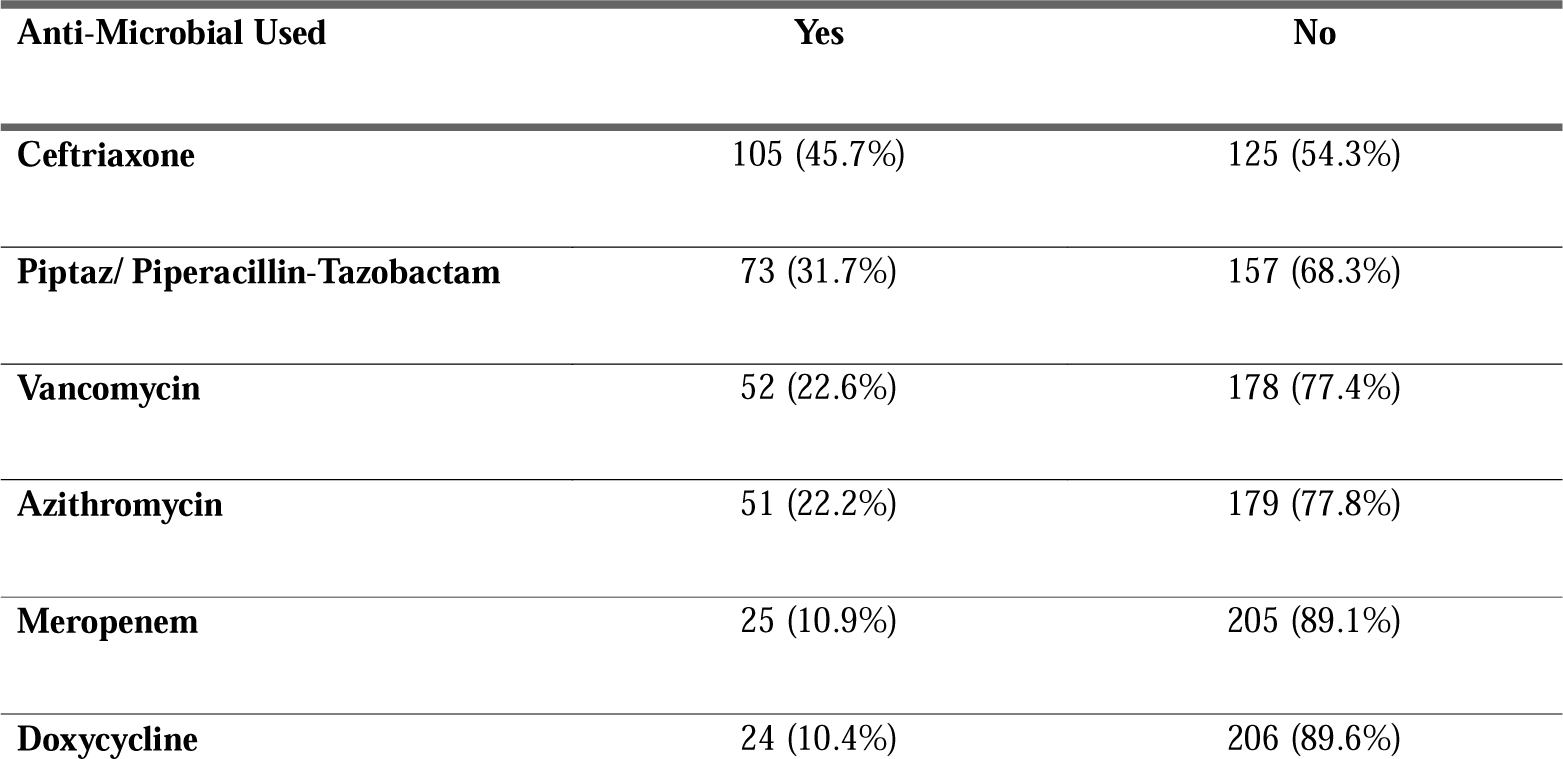
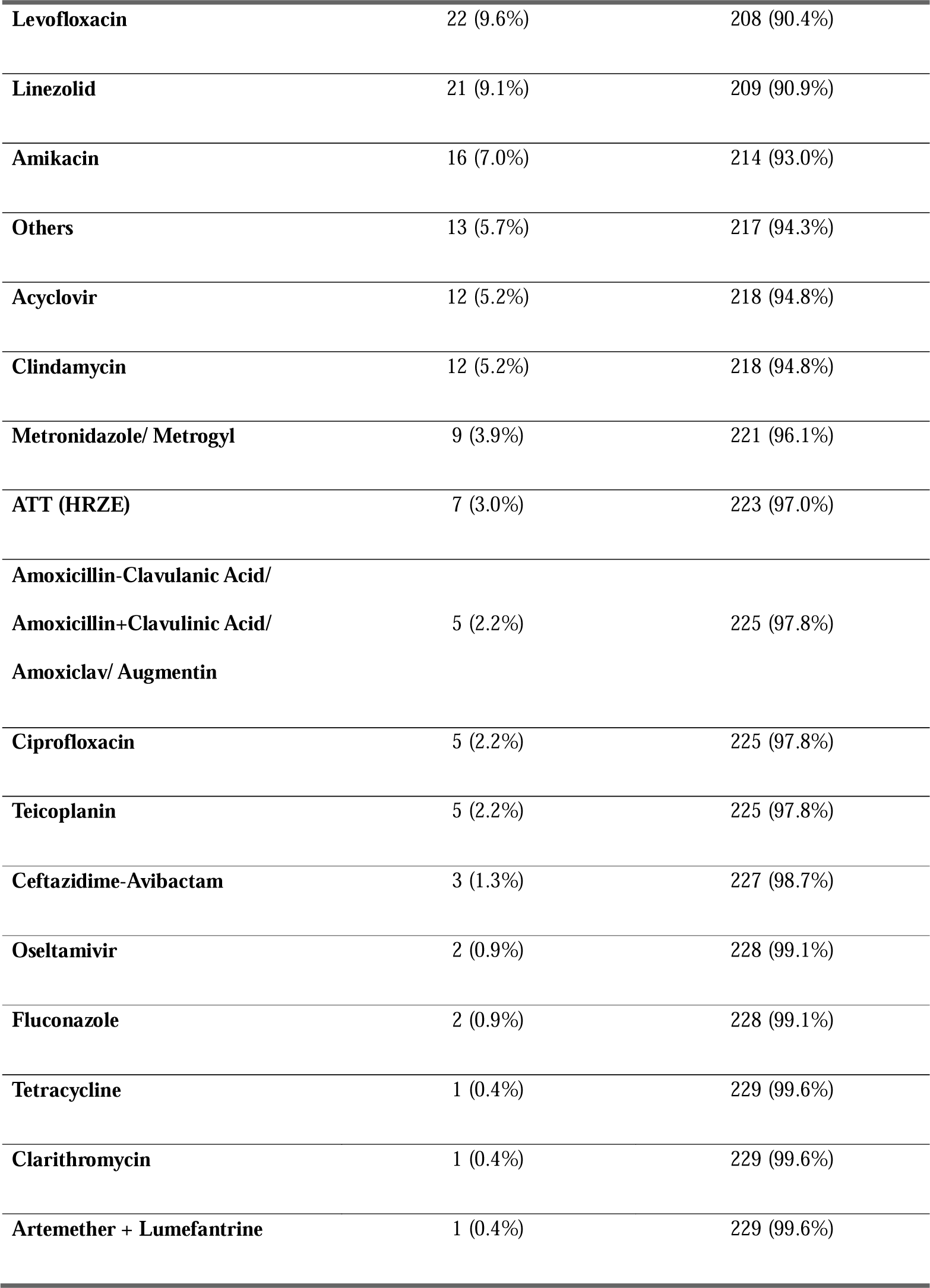
Summary of Anti-Microbial Used.

**Figure-7.**
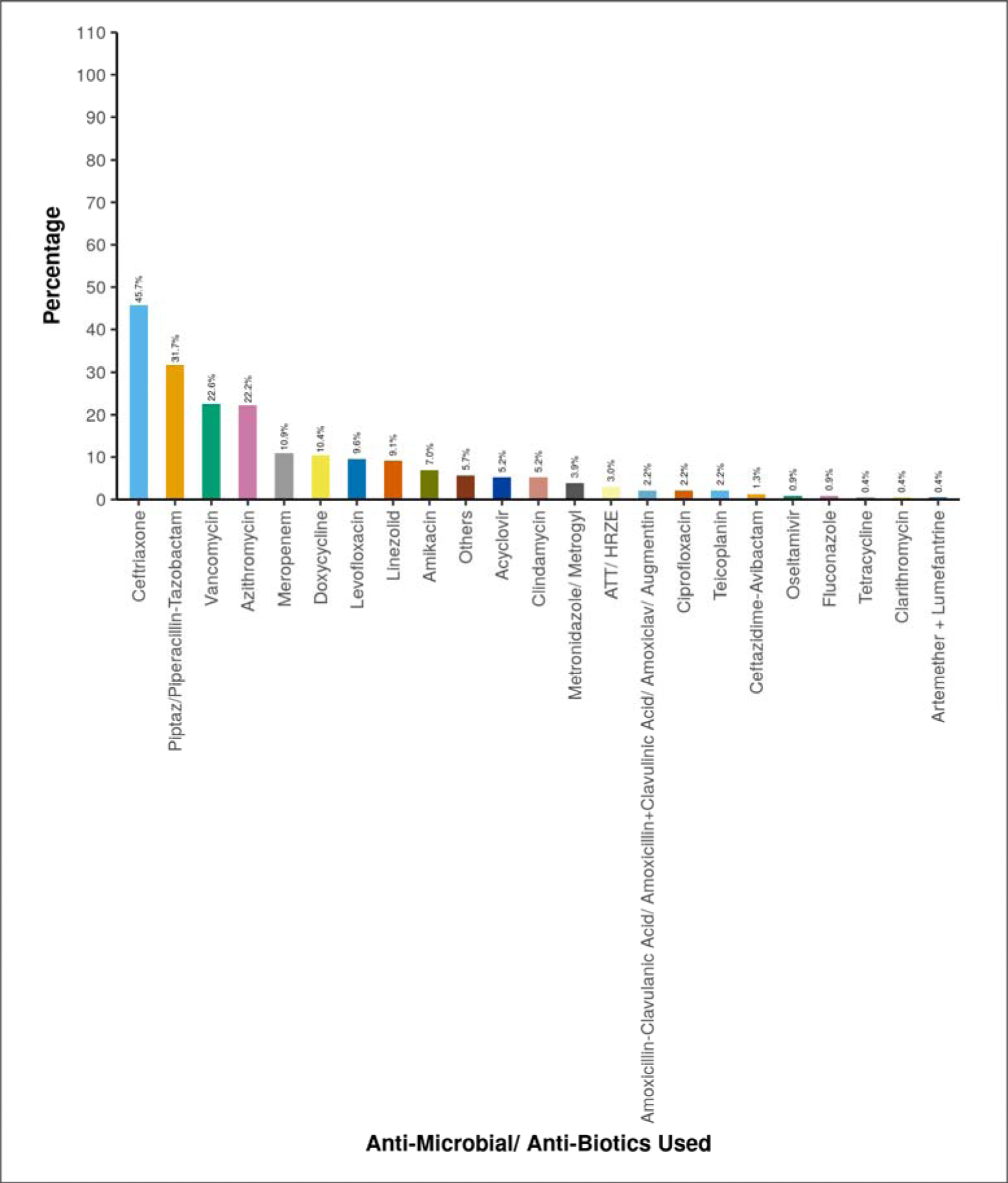
showing percentage of individual empirical antibiotics.

-Among the study cohort of 230 patients, majority of them were discharged with stable vitals (91.3%), while 6.1% of them were discharged with unstable vitals and there was an overall 2.6% mortality rate, which was equally distributed between both genders [Figure-8].

**Figure-8:**
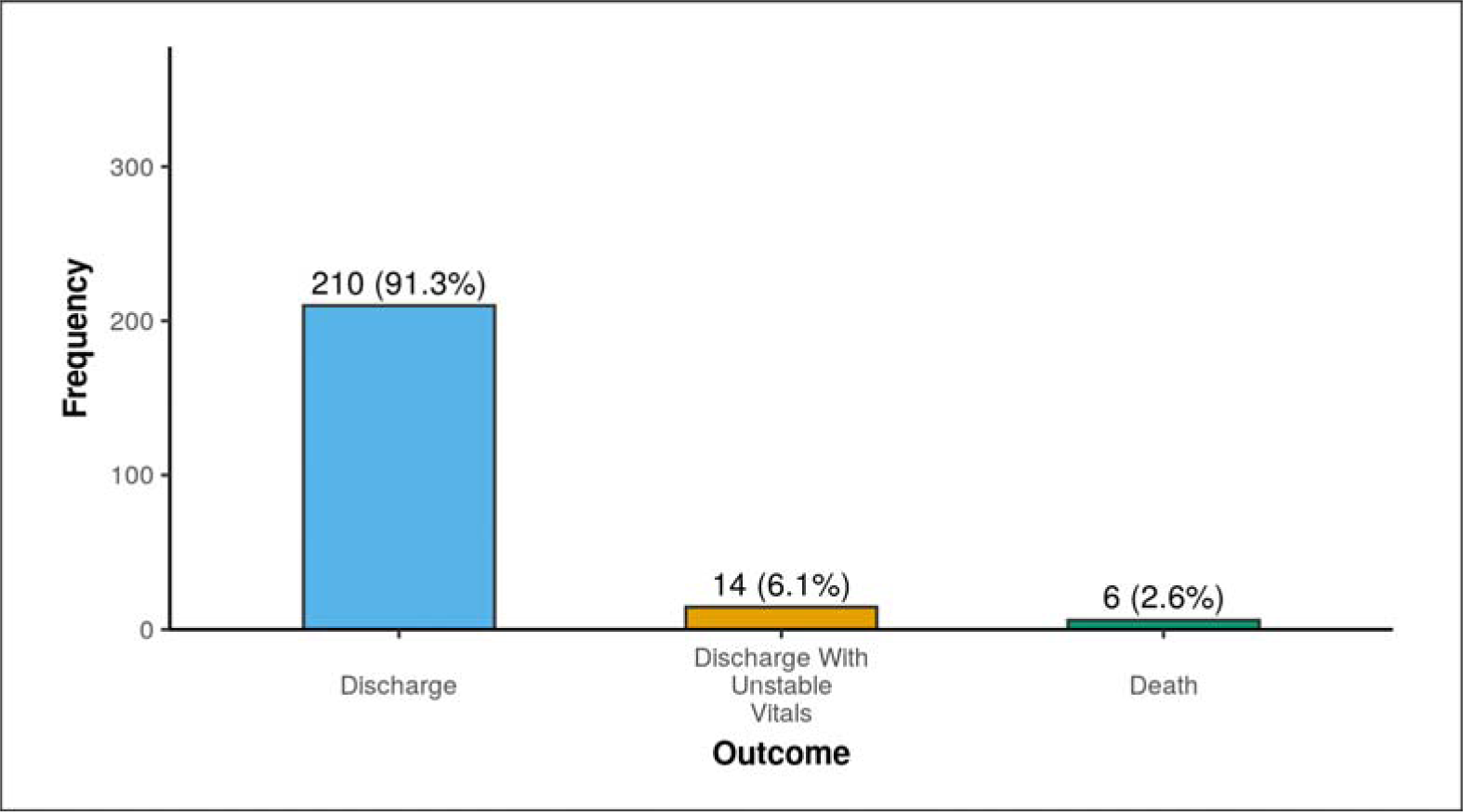
Graph showing proportion of patients’ outcome.

## DISCUSSION

This was a single centre longitudinal observational study done at a tertiary care hospital and teaching institute in Northern India which involved a study cohort of 230 patients. The aim of this study was to estimate proportion on patients in different sepsis categories and also to find the empirical antibiotics use as per AWaRe classification. As per the “Third International Consensus Definitions for Sepsis and Septic Shock (Sepsis-3)” guidelines, sepsis has been defined as life-threatening organ dysfunction which is caused by a dysregulated host response to infection (4). There have been various changes in the definition of sepsis over time with the availability of newer evidences and better understanding of the condition per se, including its pathophysiology and implications. Sepsis possesses a huge economic burden for any country, whether it is developed or developing, leading to expenditure of billions of dollars for management of sepsis. It is associated with high morbidity and mortality, with higher rates of both in developing countries, despite treatment, with data suggestive of mortality rate ranging from 15 to 56% (11).

Over the course of time, many scoring systems were introduced which can better predict the outcome and risk of mortality in patients with sepsis, such as Sequential Organ Failure Assessment (SOFA) score, Acute Physiology and Chronic Health Evaluation (APACHE) score, National Early Warning Score (NEWS) and many more, however, none of these scores can diagnose sepsis per se. In fact, these scoring systems merely denote the organ dysfunction which is associated with sepsis; however, they cannot differentiate between organ dysfunction caused by sepsis or non-sepsis conditions. This led to search for potential biomarker which can be used for identification of sepsis, and many biomarkers for sepsis have been identified (12), but only one biomarker, which is procalcitonin, is been seen with some role as per the latest guidelines of Surviving Sepsis Campaign (13,14).

All these challenges associated with diagnosis of sepsis has led us to creation of various classes of sepsis utilising a self-prepared approach involving various steps. This study was aimed to know the proportion of patients with suspected sepsis in different categories of sepsis and to also assess the changes in categories of sepsis, reflecting the dynamic nature of sepsis. The study cohort 230 patients had mean age of 40.7 ± 14.49 years with slight majority of females (50.9%) as compared to males (49.1%). Patients included were categorized into different categories of sepsis on the basis of proposed classification system, both at the time of initial presentation, which is usually on the basis of treating clinician, and also at the final time, which is usually at the time of outcome (discharge/ death). It was found that the initial categorization of sepsis in patients with suspected sepsis was dominated by the probable sepsis (51.3%) and possible sepsis (35.7%); however, the final categorization was composed of the major asepsis group (57.8%), as shown in Table-2. This change in the category or class of sepsis over the different time intervals represents the dynamic nature of sepsis.

Antimicrobial Resistance (AMR) occurs when microorganisms—such as bacteria, fungi, parasites, and viruses—adapt in ways that render antimicrobial medications, like antibiotics, ineffective against them (15). It has become one of the most pressing global challenges of the 21st century, driven by the rapid increase in resistant organisms and infections, and the shortage of new or upcoming antimicrobial drugs to address the problem (16). AMR is often called the “Silent Pandemic” and requires immediate and effective action. It should be treated as a current crisis, not a future concern (17). Without preventive measures, AMR could potentially become the leading cause of death globally by 2050 (18). Global estimates indicate that over 1.2 million deaths were directly attributed to AMR in 2019. If insufficient action is taken to address AMR, this number is projected to rise to around 10 million deaths annually by 2050 (18). To optimize antibiotic use, the World Health Organization (WHO) introduced a new classification system in 2017. This system categorizes antimicrobials into three groups—Access, Watch, and Reserve (AWaRe)—based on their spectrum, the anticipated risk of resistance development, toxicity risk, and clinical utility (19). This classification of antimicrobials has been revised twice, in 2019 and 2021 respectively (20,21). This study also estimated the use of empirical antibiotics as per WHO AWaRe classification and also the most frequently used empirical antibiotics. It was revealed from the results of the study that

The study also aimed to study the antibiotics usage in sepsis as per the WHO AWaRe (Access Watch Reserve) classification. In this study, apart from classification of patients into different sepsis categories, we also looked at the use empirical antibiotics as per WHO AWaRe classification of antibiotics and found that all included patients received empirical antibiotics. Among the total empirical antibiotics used, 26.4% of patients received antibiotics belonging to “Access” class of AWaRe classification, 92.5% of patients received antibiotics belonging to “Watch” class of AWaRe classification and 9.3% of patients received antibiotics belonging to “Reserve” class of AWaRe classification. There are studies done in various parts of the world regarding the usage of antibiotics as per WHO AWaRe classification of antibiotics. There are evidences of irrational antibiotics prescription in hospitals (22) which needs to be controlled with the formulation of standard treatment guidelines. There is still lot of knowledge gaps present regarding the AWaRe classification of antibiotics, and it has been demonstrated in a study that majority of well-trained and educated people responsible for dispensing the medications still doesn’t have proper knowledge regarding this classification of antibiotics and how it can be improved. Providing proper knowledge regarding the same has led to better selection of antibiotics when analysed in a pre and post-test manner (23). Activities of such kind should be encouraged to prevent the ongoing antimicrobial resistance. The majority of antibiotics used in our study belonged to the “Watch” group of AWaRe classification of antibiotics, which accounts for approximately 92.5%, which is way behind the WHO’s proposed target of using at least 60% of all consumed antibiotics to be from “Access” group (24).

Most commonly used empirical antibiotics in the patients involved in the study were ceftriaxone (45.7%), piperacillin-tazobactum (31.7%), vancomycin (22.6%), followed by azithromycin (22.2%), meropenem (10.9%), doxycycline (10.4%), levofloxacin (9.6%), linezolid (9.1%), amikacin (7.0%) and followed by others as shown. The AWaRe system of classification was introduced in 2017 by WHO, as a part of its antimicrobial stewardship (25), and is considered as a milestone step in the fight against the increasing antimicrobial resistance, as it was more objective in nature and a user-friendly tool for better organization of antibiotics (26). The misuse of antibiotics is a widespread issue, affecting 30% to 50% of all prescriptions. This includes instances where antibiotics are used unnecessarily, the incorrect antibiotic is chosen, or the dosage, duration, and method of administration are incorrect (27,28). The use of empirical antibiotic treatment in patients suspected of having sepsis can be justified due to the following reasons:

- **Initial Evaluation Challenges**: At the time of initial clinical evaluation, it is often difficult to accurately predict or identify which patients with sepsis will deteriorate. Sepsis can present with a wide spectrum of severity, and early symptoms can be nonspecific. This makes it challenging for clinicians to determine the exact prognosis based on initial presentation.
- **Early Phase of Sepsis:** Patients might be in the early stages of sepsis, where signs and symptoms are not fully developed, making it harder to assess the severity. During this phase, the clinical picture can be misleading, and patients who appear relatively stable may rapidly deteriorate.
- **Empirical Antibiotic Treatment:** Given the high stakes of sepsis, where delays in antibiotic treatment are associated with increased morbidity and mortality, clinicians often opt to start empirical antibiotics. This is a precautionary approach to ensure that treatment is not delayed, which could otherwise result in worse outcomes.
- **Risk of Delayed Treatment:** Evidence shows that delays in the initiation of appropriate antibiotic therapy are linked to poor outcomes in sepsis patients. Therefore, the empirical use of antibiotics is a standard practice to mitigate this risk, ensuring that patients receive timely treatment even if there is uncertainty in the initial diagnosis.

These factors underscore the necessity of empirical antibiotic therapy in the management of suspected sepsis. They also highlight the importance of continuous monitoring and reassessment to adjust treatment as more information becomes available through diagnostic tests and clinical observation.

### Limitations

Any research study done is bound to have limitations, and this one is a no exception from this rule. Despite all measures, there are associated limitations with this study which highlighted below.

⍰ **Single-Centre Setting**: This study was conducted at a single tertiary care centre and conducting the study at a single centre may limit the generalizability of the findings to a broader population. The results might reflect specific characteristics of the study site or patient population, reducing the external validity and potentially affecting the achievement of the research aims.
⍰ **Use of a novel classification approach**: The novel 3-step approach/ model prepared for this study was not previously utilised or studied in any study, making its application in real world scenario can be challenging.
⍰ **Unavailability of reference study**: Utilisation of a newly self-prepared approach was used in this study with no prior available reference study for efficacy of the approach adds to its limitations.
⍰ **External validity**: Findings from the study may not be applicable to populations with different demographic characteristics or healthcare practices, as it was conducted in a specific tertiary care centre. Therefore, the external validity and generalizability of the results to other settings need to be carefully considered.

### Interpretation

This study highlights the importance of availability of a structured approach to sepsis identification and classification, especially in resource-limited settings including India. The novel 3-step model which combines NEWS-2 score, clinical diagnosis, risk factors and available laboratory investigations provides a practical framework for clinicians to identify sepsis timely and accurately was utilised for classification of patients into different sepsis categories which classified the majority of patients into probable sepsis group (51.3%) initially, which was later dominated by asepsis group (57.8%) on final evaluation.

Empirical antibiotic usage was also studied and was classified as per WHO AWaRe (Access Watch Reserve) classification. The study revealed that 92.5% of participants received empirical antibiotics belonging to “Watch” group of classification, followed by 26.4% of participants receiving “Access” group of antibiotics, which is way behind the WHO’s proposed target of using at least 60% of all consumed antibiotics to be from “Access” group (24). Among the individual antibiotics, ceftriaxone (45.7%) was the most commonly used empirically, followed by piperacillin-tazobactam (31.7%), vancomycin (22.6%) and azithromycin (22.2%), and so on.

Inappropriate use of antibiotics without proper diagnosis and indication has led to a rampant increase in antimicrobial resistance (AMR) over the last few decades without the discovery of new antibiotics or classes of antibiotics. This study highlights the importance of right sepsis classification, as this is the need of the hour to tackle the growing burden of antimicrobial resistance (AMR). All healthcare professionals including doctors and nursing staff involved in the management of patients with sepsis should know that it is a dynamic process and similarly, for its management, use of antibiotics should be judicious. We should try to use antibiotics as per WHO AWaRe system to curb the ongoing risk of generation of new “superbugs”. Future studies are needed to improve the identification of sepsis and to classify them into categories which can be used to guide the management.

### Generalisability

As previously discussed in the limitation section, findings from the study may not apply to populations with different demographic characteristics or healthcare practices, as it was conducted in a specific tertiary care center. Therefore, the external validity and generalizability of the results to other settings need to be carefully considered and can be commented on with confidence only after a study with larger sample size and including participants from different demographic backgrounds.

## Data Availability

All data produced in the present study are available upon reasonable request to the authors

## REFERENCES

1. Torio CM, Moore BJ. National inpatient hospital costs: the most expensive conditions by payer, 2013. Healthcare cost and utilization project (HCUP) statistical briefs [Internet]. 2006 Feb.

2. Rudd KE, Johnson SC, Agesa KM, Shackelford KA, Tsoi D, Kievlan DR, Colombara DV, Ikuta KS, Kissoon N, Finfer S, Fleischmann-Struzek C. Global, regional, and national sepsis incidence and mortality, 1990–2017: analysis for the Global Burden of Disease Study. The Lancet. 2020 Jan 18;395(10219):200-11.

3. Jeganathan N. Burden of sepsis in India. Chest. 2022 Jun 1;161(6):1438–9.

4. Singer M, Deutschman CS, Seymour CW, Shankar-Hari M, Annane D, Bauer M, Bellomo R, Bernard GR, Chiche JD, Coopersmith CM, Hotchkiss RS. The third international consensus definitions for sepsis and septic shock (Sepsis-3). Jama. 2016 Feb 23;315(8):801–10.

5. Kellum JA, Formeck CL, Kernan KF, Gómez H, Carcillo JA. Subtypes and mimics of sepsis. Critical Care Clinics. 2022 Apr 1;38(2):195–211.

6. AWaRe classification of antibiotics for evaluation and monitoring of use, 2023 [Internet]. www.who.int. Available from: https://www.who.int/publications/i/item/WHO-MHP-HPS-EML-2023.04

7. Ikuta KS, Swetschinski LR, Aguilar GR, Sharara F, Mestrovic T, Gray AP, Weaver ND, Wool EE, Han C, Hayoon AG, Aali A. Global mortality associated with 33 bacterial pathogens in 2019: a systematic analysis for the Global Burden of Disease Study 2019. The Lancet. 2022 Dec 17;400(10369):2221–48.

8. Davies J, Davies D. Origins and evolution of antibiotic resistance. Microbiology and molecular biology reviews. 2010 Sep;74(3):417–33.

9. Fleming-Dutra KE, Hersh AL, Shapiro DJ, Bartoces M, Enns EA, File TM, Finkelstein JA, Gerber JS, Hyun DY, Linder JA, Lynfield R. Prevalence of inappropriate antibiotic prescriptions among US ambulatory care visits, 2010-2011. Jama. 2016 May 3;315(17):1864-73.

10. Zhao H, Wei L, Li H, Zhang M, Cao B, Bian J, Zhan S. Appropriateness of antibiotic prescriptions in ambulatory care in China: a nationwide descriptive database study. The Lancet Infectious Diseases. 2021 Jun 1;21(6):847–57.

11. Bauer M, Gerlach H, Vogelmann T, Preissing F, Stiefel J, Adam D. Mortality in sepsis and septic shock in Europe, North America and Australia between 2009 and 2019—results from a systematic review and meta-analysis. Critical Care. 2020 Dec;24:1-9.

12. Van Engelen TS, Wiersinga WJ, Scicluna BP, van der Poll T. Biomarkers in sepsis. Critical care clinics. 2018 Jan 1;34(1):139–52.

13. Pierrakos C, Vincent JL. Sepsis biomarkers: a review. Critical care. 2010 Feb;14:1–8.

14. Bloos F., Reinhart K.: Rapid diagnosis of sepsis. Virulence 2014; 5: pp. 154–160

15. World Health Organization. Antimicrobial Resistance [Internet]. World Health Organization. WHO; 2023. Available from: https://www.who.int/news-room/fact-sheets/detail/antimicrobial-resistance.

16. Prestinaci F, Pezzotti P, Pantosti A. Antimicrobial resistance: a global multifaceted phenomenon. Pathogens and global health. 2015 Oct 3;109(7):309–18.

17. Founou RC, Blocker AJ, Noubom M, Tsayem C, Choukem SP, Dongen MV, Founou LL. The COVID-19 pandemic: A threat to antimicrobial resistance containment. Future Science OA. 2021 Sep;7(8):FSO736.

18. O’Neill J. Review on antimicrobial resistance: tackling drug-resistant infections globally: final report and recommendations. Review on antimicrobial resistance: tackling drug-resistant infections globally: final report and recommendations. 2016.

19. World Health Organization. The selection and use of essential medicines: report of the WHO Expert Committee, 2017 (including the 20th WHO Model List of Essential Medicines and the 6th WHO Model List of Essential Medicines for Children). Geneva, Switzerland: World Health Organization; 2017.

20. Sharland M, Gandra S, Huttner B, Moja L, Pulcini C, Zeng M, Mendelson M, Cappello B, Cooke G, Magrini N, Aziz Z. Encouraging AWaRe-ness and discouraging inappropriate antibiotic use—the new 2019 Essential Medicines List becomes a global antibiotic stewardship tool. The Lancet Infectious Diseases. 2019 Dec 1;19(12):1278–80.

21. The selection and use of essential medicines. World Health Organization; 2022.

22. Mugada V, Mahato V, Andhavaram D, Vajhala SM. Evaluation of prescribing patterns of antibiotics using selected indicators for antimicrobial use in hospitals and the access, watch, reserve (AWaRe) classification by the World Health Organization. Turkish Journal of Pharmaceutical Sciences. 2021 Jun;18(3):282.

23. Abu-Ajaleh S, Darwish Elhajji F, Al-Bsoul S, Abu Farha R, Al-Hammouri F, Amer A, Al Rusasi A, Al-Azzam S, Araydah M, Aldeyab MA. An Evaluation of the Impact of Increasing the Awareness of the WHO Access, Watch, and Reserve (AWaRe) Antibiotics Classification on Knowledge, Attitudes, and Hospital Antibiotic Prescribing Practices. Antibiotics. 2023 May 23;12(6):951.

24. Zanichelli V, Sharland M, Cappello B, Moja L, Getahun H, Pessoa-Silva C, Sati H, van Weezenbeek C, Balkhy H, Simão M, Gandra S. The WHO AWaRe (Access, Watch, Reserve) antibiotic book and prevention of antimicrobial resistance. Bulletin of the World Health Organization. 2023 Apr 4;101(4):290.

25. Sharland M, Zanichelli V, Ombajo LA, Bazira J, Cappello B, Chitatanga R, Chuki P, Gandra S, Getahun H, Harbarth S, Loeb M. The WHO essential medicines list AWaRe book: From a list to a quality improvement system. Clinical Microbiology and Infection. 2022 Dec 1;28(12):1533–5.

26. Mudenda S, Daka V, Matafwali SK. World Health Organization AWaRe framework for antibiotic stewardship: Where are we now and where do we need to go? An expert viewpoint. Antimicrobial Stewardship & Healthcare Epidemiology. 2023;3(1):e84.

27. Fleming-Dutra KE, Hersh AL, Shapiro DJ, Bartoces M, Enns EA, File TM, Finkelstein JA, Gerber JS, Hyun DY, Linder JA, Lynfield R. Prevalence of inappropriate antibiotic prescriptions among US ambulatory care visits, 2010-2011. Jama. 2016 May 3;315(17):1864-73.

28. Zhao H, Wei L, Li H, Zhang M, Cao B, Bian J, Zhan S. Appropriateness of antibiotic prescriptions in ambulatory care in China: a nationwide descriptive database study. The Lancet Infectious Diseases. 2021 Jun 1;21(6):847–57.

